# Community co-designed workshops build confidence in use of functional evidence for variant classification

**DOI:** 10.1101/2025.11.10.25339935

**Authors:** Rehan Marie Villani, Bronwyn Terrill, Emma Tudini, Ebony Matotek, Tessa Mattiske, Corrina C. Cliffe, Bryony A. Thompson, Laveniya Satgunaseelan, Christopher N. Hahn, Ben Lundie, John Christodoulou, Alan F. Rubin, Amanda B. Spurdle

## Abstract

**Purpose:** To increase diagnostic yield from genomic testing, it is essential to improve the use of experimental data as functional evidence in variant classification. Recent work suggests building confidence of those performing functional data evaluation may improve their capability to use functional evidence. We present a solution to develop confidence and build capability while supporting practice development, through workshop-based community discussion and consultation.

**Methods:** A series of workshops were delivered, each with learning content centred on current expert recommendations while surveying participants in their experimental evaluation practice and opinions around evaluating experimental data.

**Results:** Workshop participants were highly engaged and indicated a desire for further training and resources. Consultation suggested that ambiguity in the current recommendations underpinned differences in interpretation and ultimately resulted in practice variation. Collaborative, case-based discussion was identified as the overall preferred method of building confidence, so we propose this as an approach to field-based capability development. We delivered case-based community discussion, after which participants reported improved comfort in the evaluation practice.

**Conclusion:** Through the consultative workshop approach, workshop participants have built confidence but also contributed to developing field recommendations. Therefore, we recommend this approach as a means to capability development for evolving fields.

## Introduction

In 2015 a landmark paper by the American College of Medical Genetics and Genomics (ACMG) and Association for Molecular Pathology (AMP) ^1^ described guidelines for variant classification. The guidelines defined a process for evaluating the clinical significance of a genetic variant by assessing all available evidence to determine if the genetic variant is pathogenic (disease-causing) or benign. Along with a number of additional recommendations and clarifications, the guidelines are now used worldwide ^2^.

Additional recommendations for application of functional evidence define how to evaluate the suitability and strength of evidence for experimental data. They dictate that experimental assay results must first be rigorously assessed to determine if the assay is an appropriate model and assay type ^3^. Then, if the assay is of appropriate quality, the applicable strength of evidence for curation can be determined. The recommendations state that functional evidence, if determined to be sufficiently predictive, can be applied at up to strong evidence (PS3 or BS3 codes, following current 2015 ACMG/AMP guidelines). Subsequent alignment of the ACMG/AMP guidelines to a Bayesian framework defined an approach to calculating the predictive power for evidence types ^4^. Application of these predictive calculations to functional studies have demonstrated the value of higher-throughput functional evidence for classification ^3^, and has shown the evidence has sufficient weight to impact the variant classification tier, particularly when applicable at Strong evidence level ^5,6^. Specific recommendations have also been provided to guide the generation of data using high-throughput approaches ^7^.

The AusMAVE Education project was established to improve functional evidence use and encourage the adoption of Multiplexed Assays of Variant Effect (MAVEs) for variant interpretation. Early project outcomes ^8^, along with other international studies (CanVIG and AVE) ^9; 10^, have shown that even expert variant curators are not confident in the assessment of functional evidence for clinical curation. This observation was supported by empirical findings from Shariant, the Australian and New Zealand laboratory clinical variant classification results sharing platform ^11^: functional evidence was one of the least applied evidence types in Shariant and there were often differences between laboratories in the application of evidence for the same variant. In addition, analysis of both Shariant and ClinVar information showed that functional evidence was a significant cause of variant classification discordance between laboratories (https://shariant.org.au) ^12^. Combined, these observations indicated there is still work to do to clarify difficult aspects of functional evidence evaluation across the breadth of functional assay types.

To improve the use of functional evidence in variant classification, the AusMAVE Education Team developed a series of workshops centred on evaluating functional assays. The workshops demonstrated practice using a case-based approach, but also investigated current practice and assessed participant preference of expert-identified solutions. Here we present the materials and learnings from the workshops, and propose a model to support capability building in an evolving practice such as variant classification.

## Methods

### Planning and establishment

An advisory committee (the AusMAVE Education Working Group) composed of field experts was established at the outset of the project. The nascent committee first identified crucial stakeholder groups and then recruited representative members of identified groups. Stakeholders identified included professional bodies such as the Human Genetics Society of Australasia (HGSA), the Royal College of Pathologists of Australasia (RCPA) and the Australian Functional Genomics Network (AFGN), as well as generators and users of functional data including research scientists, clinical scientists, bioinformaticians and clinical users of variant testing results. Working group membership was reassessed mid-term to ensure continuing representation of appropriate stakeholders.

A scoping review was performed at the outset of the project and a living program plan developed to define the objectives (project documents are hosted on Zenodo and available via the AusMAVE project page, https://www.qimrb.edu.au/ausmave). These documents were considered by the advisory committee to determine the initial approach and define activities. The advisory committee guided establishment of learning and research objectives, interpretation of results, and development and review of outcomes and deliverables. A pre- and post-implementation evaluation approach was identified early^13^, and developed throughout the lifetime of the project.

Participants were invited to join project activities through the identified stakeholder groups. The research undertaken in this study was approved by the Human Research Ethics Committee of QIMR Berghofer, Project ID P3920.

### Workshops

Three interactive workshops were held between November 2023 and November 2024 (see Supplemental Information; Program Summary). Both the workshops and the overall program design were aligned with the Australian Genomics Education Program Logic model ^14^. Workshop 1 was delivered in-person, in association with the Australasian Society of Diagnostic Genomics Special Interest Group Meeting. Workshop 2 and Workshop 3 were delivered online with recruitment through the Human Genetics Society of Australasia, the Australian Functional Genomics Network and Shariant. A member contact list was also established, enabling existing members to invite additional participants through their local network across the life of the program.

Our previous work ^8^ identified that some of the major barriers to functional evidence use include a lack of familiarity with experimental assays, difficulty finding data, and lack of training. Workshop activities delivered content related to the evaluation of functional assays for use as functional evidence in variant classification. We defined functional evidence evaluation to comprise the aspects of assessing functional data as denoted in Brnich et al. 2019 ^3^, combined with the evidence strength calculation process described in Tavtigian et al. 2018 ^4^. Each activity contained training content aligned with the specific activity objectives and a series of consultative questions, the answers of which were analysed and used to develop the subsequent workshop objectives and associated resources (Figure 1).

**Figure 1.**
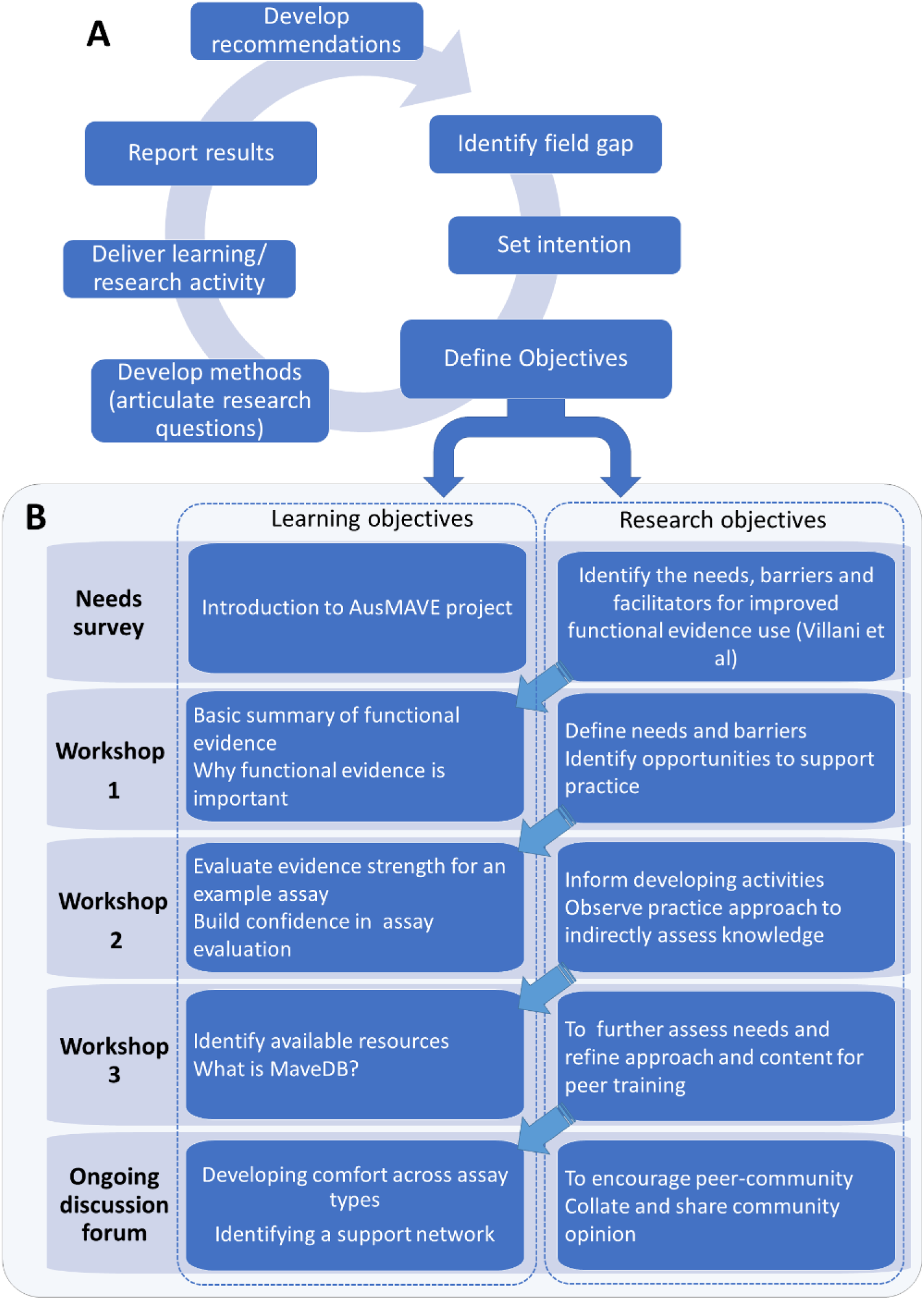
Program approach and overview. A) Summary of the iterative design approach used to develop each activity within the program. B) Summary of learning objectives (intended objectives for the workshop participants) and research objectives (intended objectives of incorporated survey) for each activity. The research results were used to inform the subsequent learning objectives. These methods are explained in more detail in Table 1, and in Supplemental information.

**Table 1.**
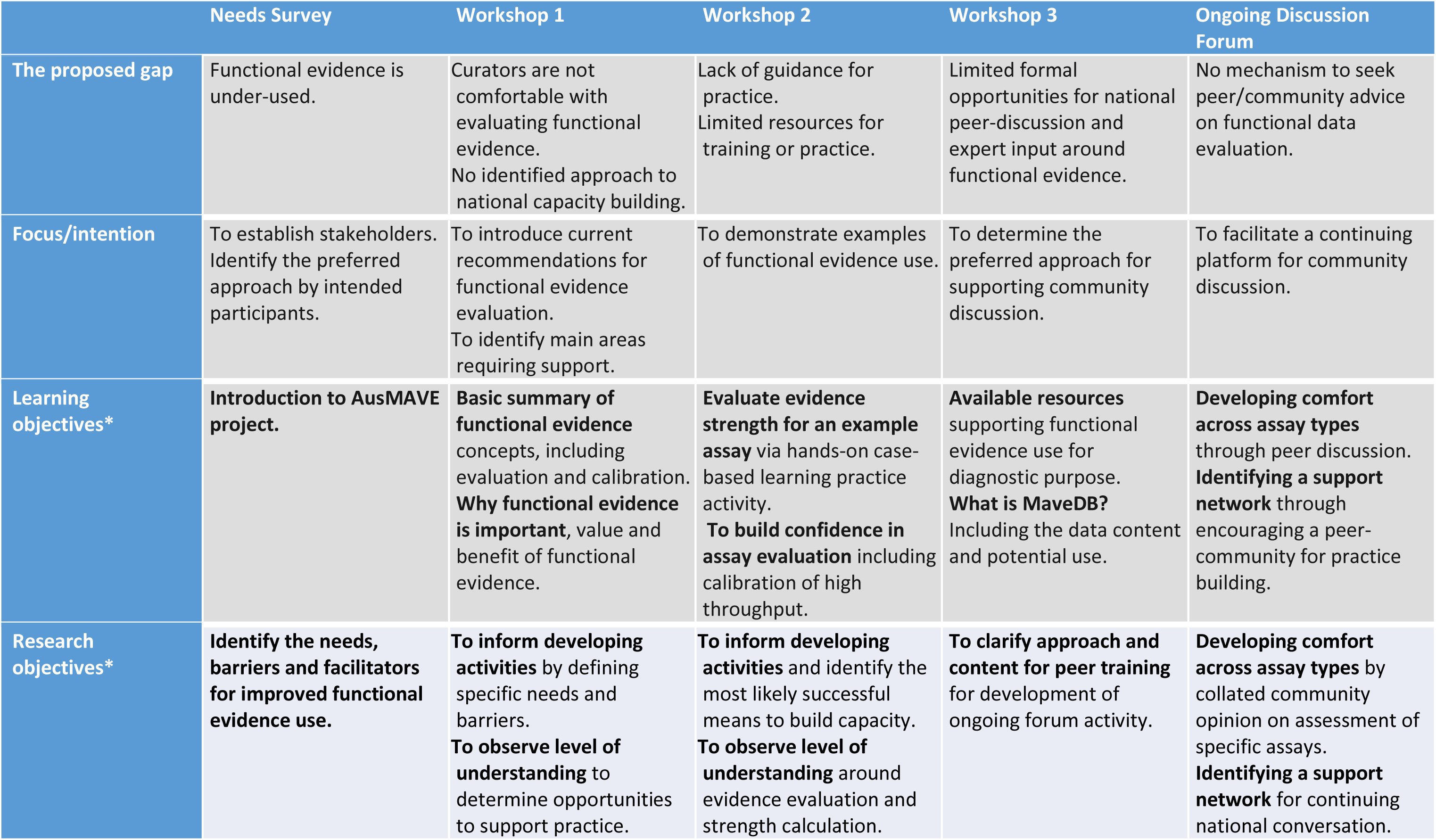
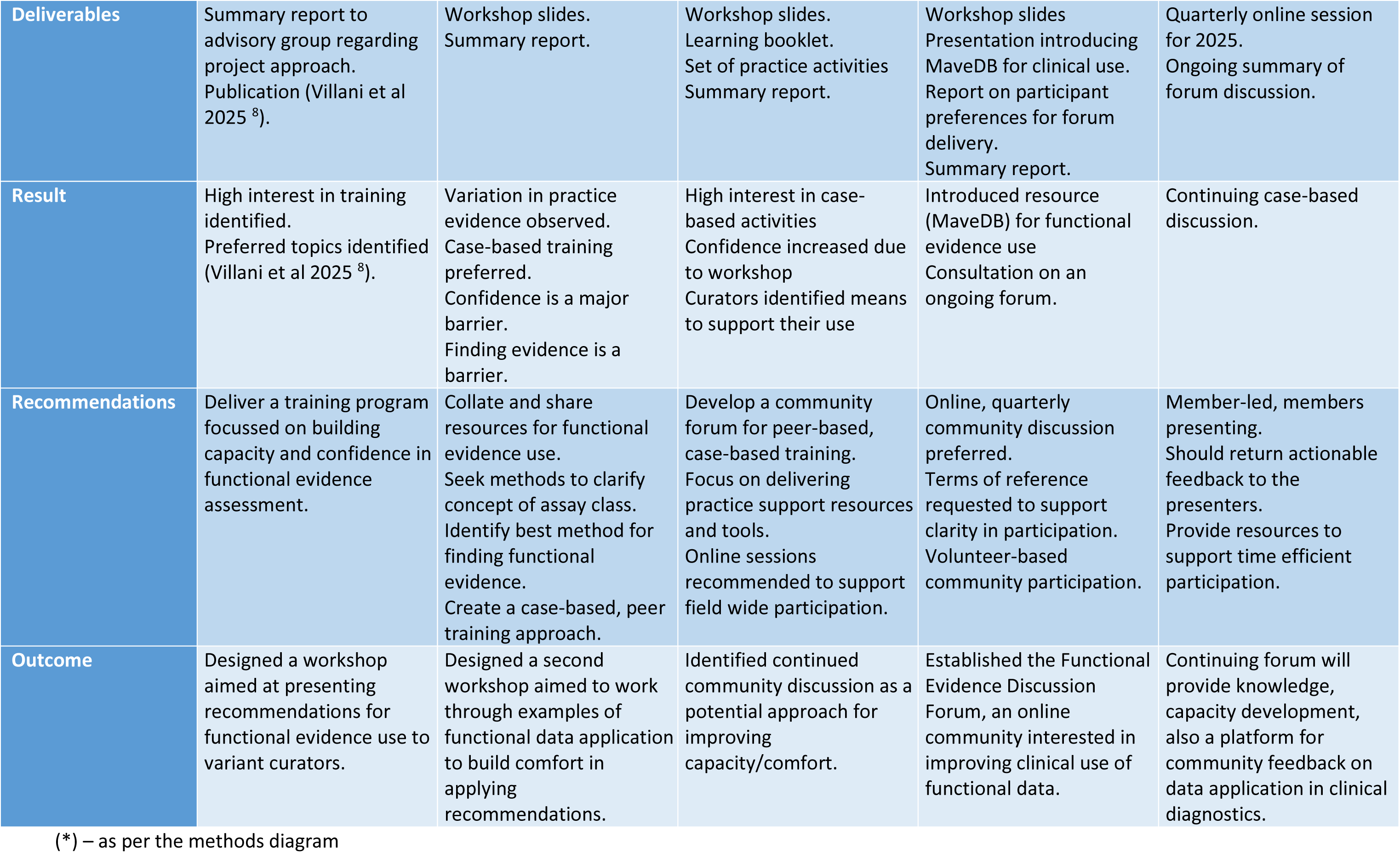
Summary of AusMAVE Education Program.

### Consultation and Surveys

Field consultation was performed through surveying the workshop and activity participants. Participants were invited to participate in the program activities through open invitation distributed via professional bodies representing the identified stakeholders, including Shariant, MaveDB, the Human Genetics Society of Australasia, the Australasian Society of Diagnostic Genomics, the Australian Functional Genomics Network and the Royal Society of Pathologists Australasia. A priority audience was defined as clinical variant curators and functional genomics researchers. Participants were asked to complete surveys via an online form, the survey platform was determined based on the workshop delivery. Workshop 1 (in-person) and Workshop 2 (online) survey consultation was delivered via google forms, accessed via personal devices through QR codes presented in the workshops. Workshop 3 was delivered online and integrated Zoom polls were used to collect participant consultation data.

### Thematic barrier analysis

Theme categories were identified in the qualitative responses and supplemented with field notes taken during the workshop discussion (by Rehan Villani). After conventional qualitative content analysis, the number of times a particular category was identified was counted and compared with other categories ^15^.

### Data Analysis

R (R version 4.3.2 (2023-10-31 ucrt))^16^/RStudio (2024.4.2.764)^17^ was used to perform analyses and generate figures. This included calculating summary statistics, such as means and confidence intervals, and the various counts provided within this report.

## Results

### Workshop 1 indicated practice variation in functional evidence evaluation

The first workshop presented a summary of the functional evidence evaluation process and used a single variant (NM_007294.4:c.5246C>G, referred to as BRCA1c.5246C>G (p.P1749R)) as an example, presenting a low-throughput ^18^ and a high-throughput assay ^19^ of relevance to the variant. We then surveyed participant’s opinion on the applicable evidence for variant classification provided by each of the two assays (Figure 2A-F). Participant opinion on the applicable evidence was varied for both assays, and additionally, between ∼15-33% of participants did not answer the question, which may suggest that they were not confident enough to answer.

**Figure 2:**
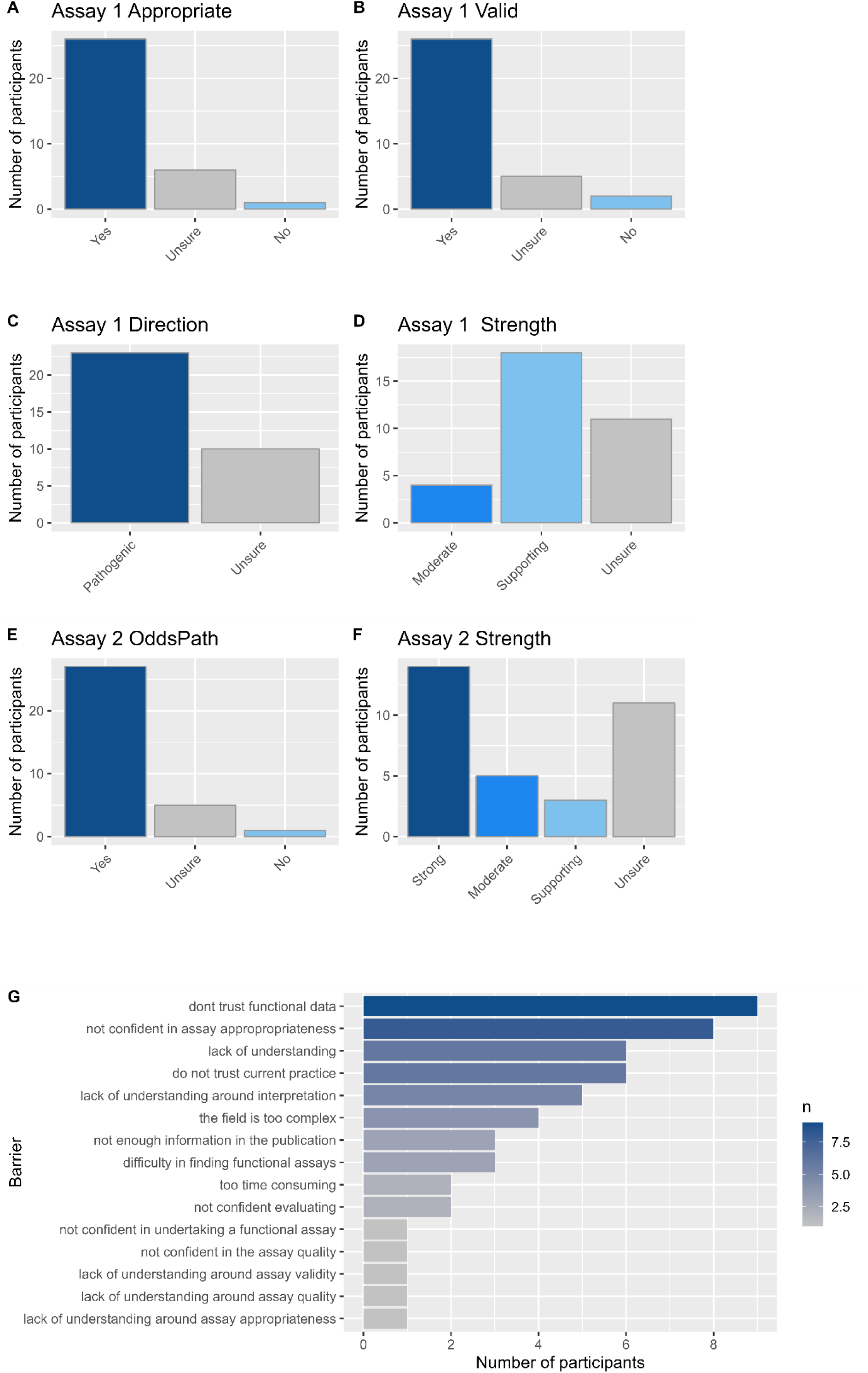
Summary results of participant opinion around functional evidence evaluation. Workshop 1 participants were shown two assay examples and asked their opinion on the assay’s use for the classification of BRCA1 variant BRCA1c.5246C>G (p.P1749R). A-D Scully et al. ^18^, and E-F Findlay et al. ^19^. A) Participant opinion on the assay as an appropriate model for disease. B) Participant opinion on the assay validity for use. C) Participant opinion on evidence direction, options given were ‘pathogenic’, ‘benign’ or ‘unsure’. D) Participant opinion on the evidence strength applicable for the assay. E) Participant opinion on whether an OddsPath/likelihood ratio should be calculated for the assay in question, Findlay et al., and F) Participant opinion on the evidence strength applicable for the assay. G) Participant opinion from free text responses on the barriers to functional evidence use. A-G, n = 33 survey participants.

During Workshop 1, we built upon results from the needs assessment ^8^ by asking participants to answer via free text ‘What do you think are barriers to functional evidence use?’. When categorised and counted, the most common barriers to functional evidence use mentioned by participants were a lack of trust in the assay data, including a lack of confidence in the assay’s quality and appropriateness for clinical use, and not fully understanding the process of functional evidence evaluation (Figure 2G).

A second workshop on Functional Evidence Evaluation was designed in consultation with the AusMAVE Education Working Group. Workshop 2 responded to the variation in practice identified in Workshop 1, by focusing on defining the functional assay evaluation process and using case example activities to demonstrate approach. Workshop 2 was delivered online; 104 participants registered, and at least 80 attended (based on the Zoom participant count, in some instances several individuals from a single site attended on one shared connection). The workshop integrated consultation by polling the participants regarding barriers to functional evidence use. Poll results identified that the majority of participants agreed that conflicting results and controls were barriers to application of specific assays as evidence in variant curation (Figure 3A): 76.6% indicated ‘there are conflicting assay results’ and 68.1% indicated ‘I am not confident the experiment has used the right controls’.

**Figure 3:**
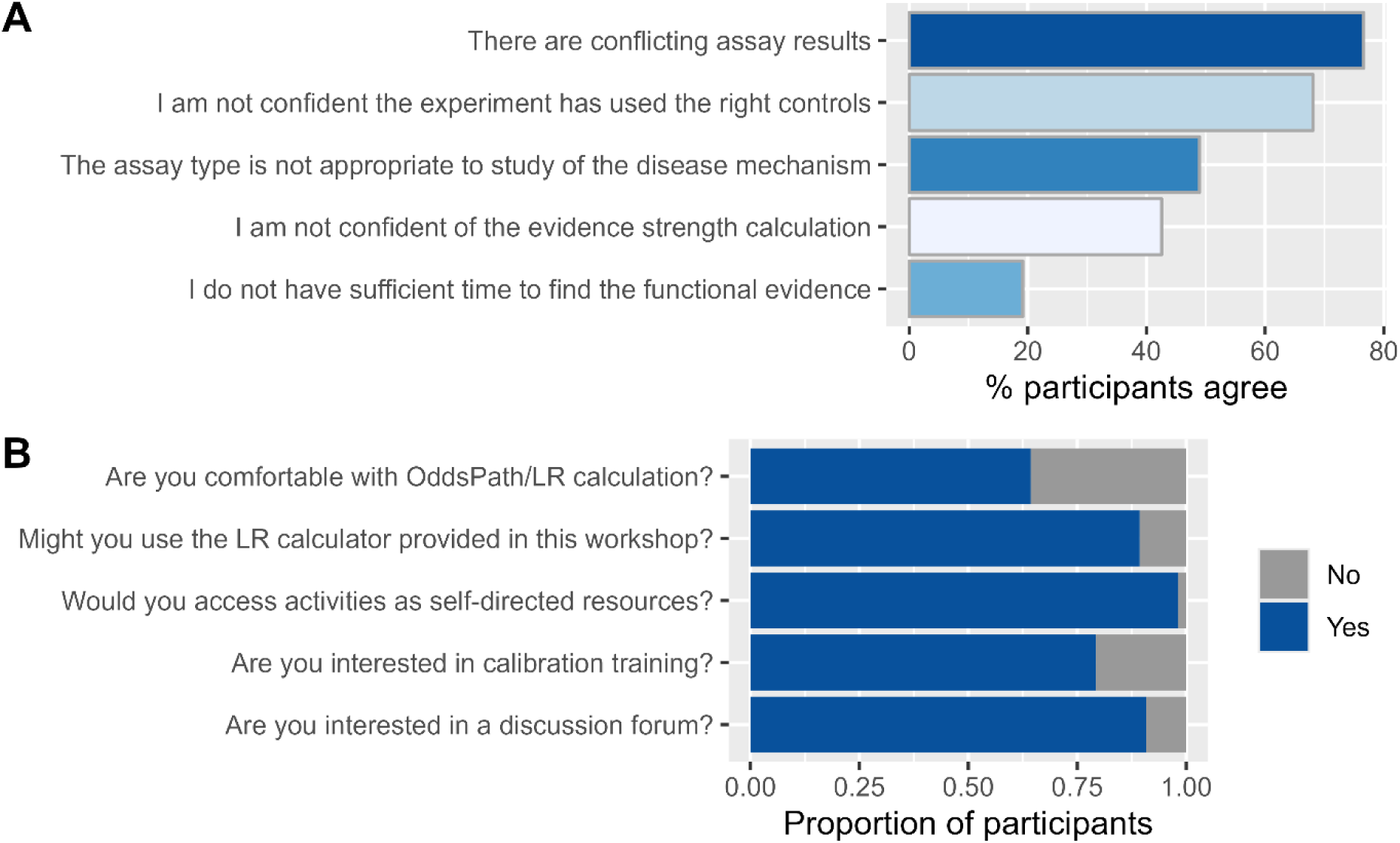
Workshop 2 consultation results. A) Participants were asked to indicate their agreement with statements that ‘explained your decision not to apply functional evidence’, agreement with each categorical statement (y axis) indicated as percentage of n = 47 total survey participants. B) Proportion of participants indicating agreement with specific statements; Are you comfortable with OddsPath/LR calculation and use of assay OddsPath/LR to determine the evidence strength of a variant? n = 47 survey participants; Do you think you may use the LR calculator provided in this workshop to support your curation? n = 57 survey participants; Would you be interested to access the workshop activities as self-directed resources? n = 57 survey participants; Would you be interested in further training on calibrating functional assays? n = 55 survey participants; Would you be interested to join a functional evidence discussion forum scheduled online meeting or online discussion page? n = 47 survey participants.

Workshop 2 demonstrated a process for calibration of functional data from a published high-throughput assay ^19^ for use as evidence in variant classification. Functional evidence strength calculation was performed based on a reference clinical control set of variants classified (or assumed) as pathogenic or benign by diagnostic labs or expert curation groups and calibration was demonstrated according to current international functional evidence recommendations ^3; 4^. Participants were asked questions about their comfort, approach to, and educational preferences for aspects of functional evidence calibration. OddsPath calculation was presented in the format of likelihood ratio (LR) estimation, investigated specifically as a central aspect of evidence strength calculation. The majority (64.3%) of participants responded that they were comfortable with using this statistical approach to estimate a likelihood ratio for or against pathogenicity for functional assay results, and thereby assign evidence strength (Figure 3B). An even larger proportion of participants (89.3%) indicated that they would use an LR calculator tool developed by the ENIGMA consortium that was provided in the workshop in practice (Figure 3B). Additionally, participants almost universally (98%) supported accessing the activities delivered in the workshop as a self-directed resource and were interested in additional training in calibrating functional assays (Figure B).

### Consultation identified overall support for a community-based collaborative approach to knowledge development

The combined consultation results from the previous Needs Survey ^8^, Workshop 1 and Workshop 2 ultimately indicated that the preferred approach to building capability would be through a community of practice approach ^20^. The consultation also indicated that discussion on case presentations or real-world scenarios could support building stakeholder confidence. In Workshop 2, participants were specifically asked to indicate their interest in attending a community discussion forum, and 49/54 (90.1%) participants indicated ‘Yes’. Due to the high level of interest identified in Workshop 2, and supported by AusMAVE Education Project Advisory Committee opinion, Workshop 3 was designed around further consultation to refine an approach to a continuing forum. Workshop 3 was delivered online and paired consultation with an information session on MaveDB, a resource that supports sharing of high-throughput functional data ^21^. Consultation integrated into the launch session explored the community’s preferred format for community discussion, with the majority of participants indicating preference for a quarterly online discussion session (Figure 4A). Around half of the participants were interested in an out-of-session chat forum (Figure 4B).

**Figure 4:**
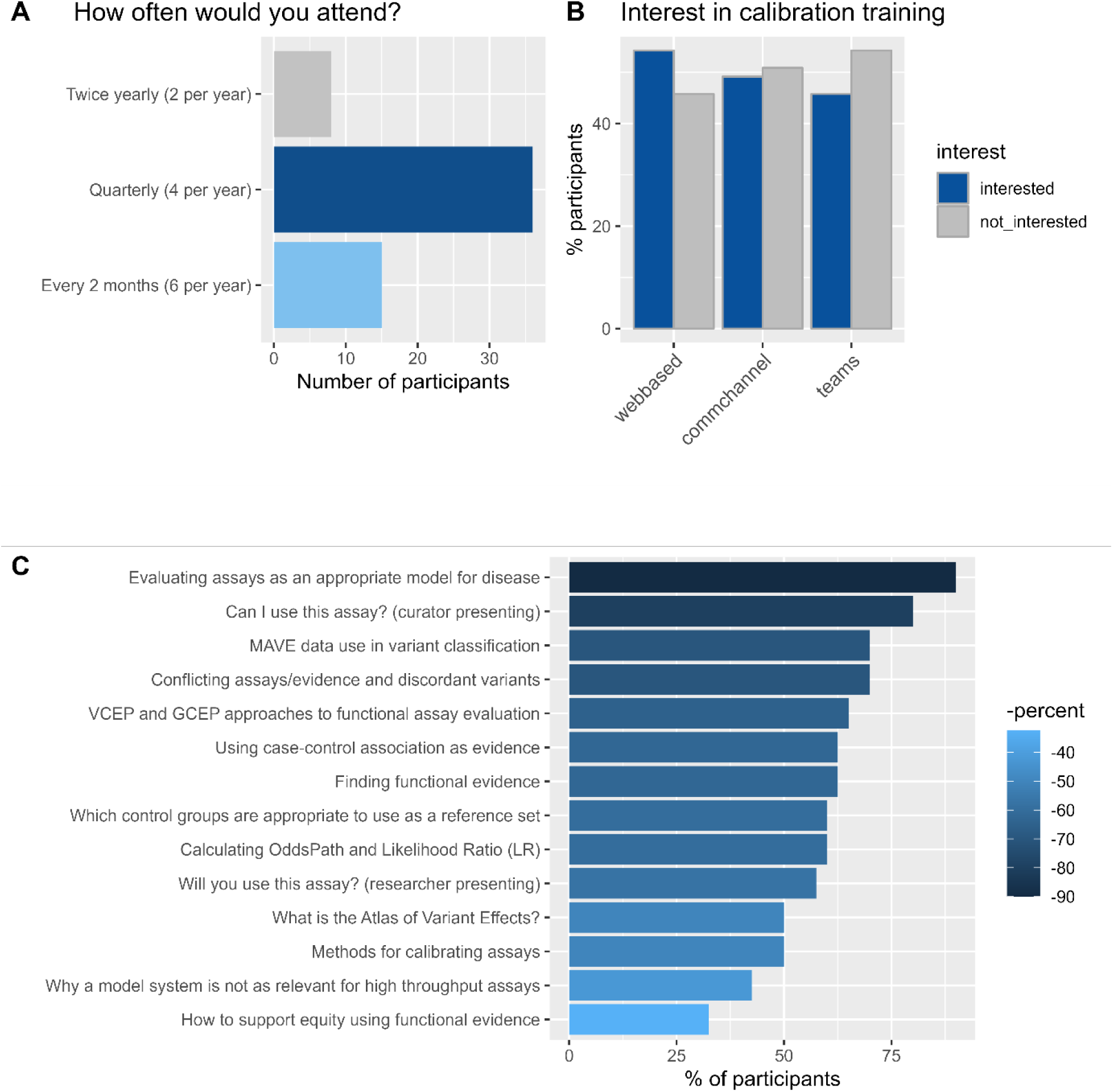
Workshop 3 determined participant preference for a community-based continuing program. A) Participants were asked to indicate their preferred regularity of sessions, from 2 per year, 4 per year, 6 per year or monthly, n = 59 respondents B) Participants were also asked regarding their preferred method of online forum (webbased), a communication channel such as a chat based discussion platform (commchanel) or via Microsoft Teams (teams), n = 59 respondents, and C) Percent of participants supporting each of the indicated topics, n = 40 respondents.

### Capacity building workshops improve confidence

Workshop participants were asked to indicate their confidence on a sliding scale at various points during the program. In an anonymous post-program assessment, 6 respondents suggested the delivered learning content was appropriate for the audience (Supplemental information). Workshop 2 was developed from expert consideration, by the AusMAVE Education Working Group, of results from the two previous consultation activities (Needs survey^8^ and Workshop 1). Participants were asked to indicate if their comfort with functional evidence application increased across the course of Workshop 2 (Figure 5A, before, compared to Figure 5B, after Workshop 2). The responsive program approach was correlated with an increasing average confidence score across activities (Figure 5C), suggesting the consultative approach is a successful means of improving confidence/comfort in functional evidence evaluation.

**Figure 5:**
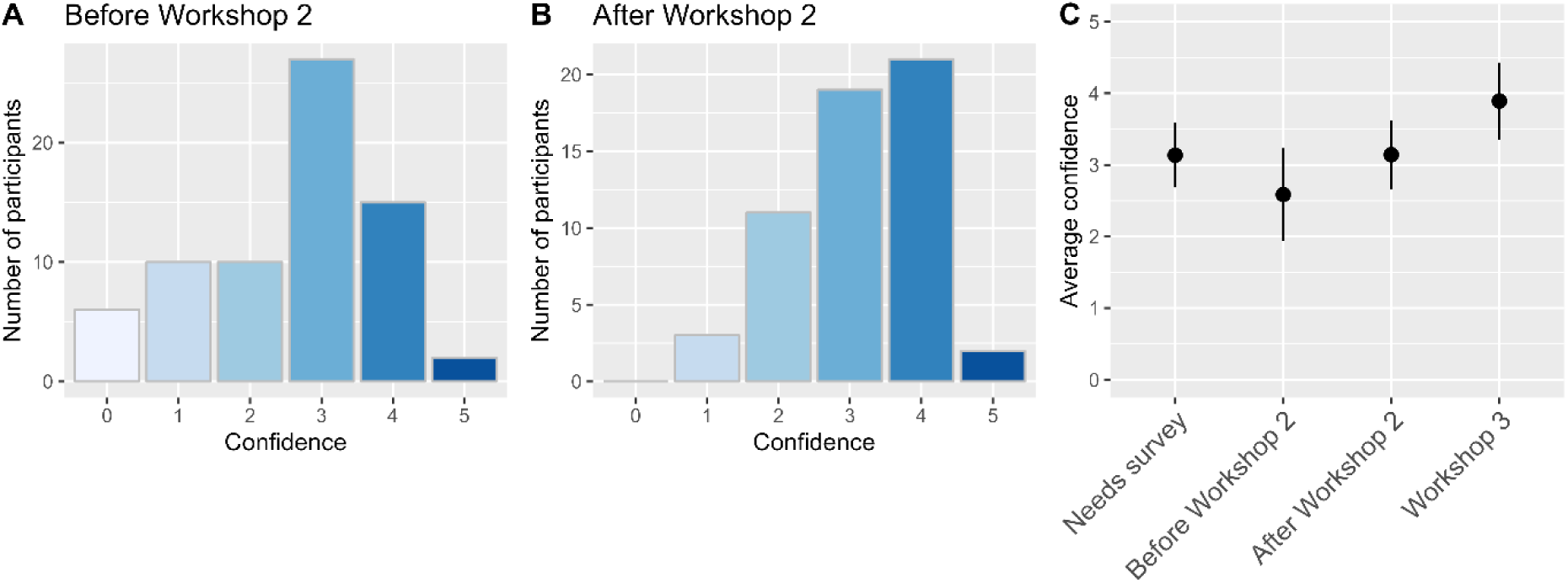
Self-reported confidence levels throughout the program. Participants were asked to indicate their confidence on a scale of 0:Not at all comfortable to 5:Very comfortable at multiple points during the program. A) Number of participants indicating each comfort levels at A) start of Workshop 2 (total n = 75) and B) end of Workshop 2 (total n = 56) C) Mean (+/- standard deviation) of comfort scores indicated at the indicated activity points through the program (Needs survey n = 33, Before workshop 2 n = 75, After workshop 2 n = 56, and during Workshop 3 n = 84).

### Community of practice identified as preferred approach to capacity building in functional evidence evaluation

The program delivered a number of capacity building activities over time, each aimed at improving clinical application of functional evidence. Along with providing learning materials, this program also consulted the participants regarding the specific areas for development and best approach for functional evidence evaluation. Table 1 summarises the overall development of objectives and outputs. Each activity identified a gap, or problem, for which we designed both educational content and aligned research questions for consultation. The consultation results then enabled the AusMAVE Education Working Group to use field opinion to inform the program direction for the next activity.

**Table 1: Co-designed diagnostic genomics training model**

## Discussion

This project demonstrates implementation of an iterative approach to capability building for translating experimental data into functional evidence for variant classification. Through integrated consultation we facilitated ongoing design and development of a program that improved stakeholder comfort with functional evidence evaluation, captured metrics to measure improvement in practice, and ultimately culminated in a community-designed Discussion Forum for continued education and practice development.

The capability building approach involved a cyclic process of: 1) identifying the practice gap; 2) presenting current understanding from the field to bridge the gap; 3) consulting to define the problem and potential solutions; 4) using the results to clarify/identify the subsequent gap. We attribute the generally positive response to the program to the comprehensive consultation at each stage. Early establishment of an expert committee, the AusMAVE Education Working Group, also proved crucial, particularly in ensuring stakeholder representation was appropriate. This membership was also reviewed and updated across the life of the program to reflect the program development. Broad recruitment of participants to the program was also prioritised to support diverse participation, along with consultation regarding the practical aspects of participation. Online activities were identified as a preference early in the consultation process by participants. This approach likely minimised barriers to attendance; clinical curators have limited time, limited budget for activities considered out of practice scope, and physical distance between cities in Australia impacts face-to-face participation in group activities. Each workshop therefore represented a concurrent learning and research activity, from which new practice tools and recommendations could be co-designed. We propose this method as a solution to concurrent capability and practice building, crucial to the developing field of variant classification. We suggest it may also be a viable model for capability building for other emerging technologies and practice.

Program consultation culminated in a community-designed output, the ‘Functional Evidence Discussion Forum’, a series of community-led discussion sessions capturing various aspects of functional evidence evaluation. Participants self-identified the activity, location, regularity and content of the forum. This forum was designed to be an online quarterly meeting to host both scientists generating assay results, and variant classification scientists applying the results to present data and raise questions, thereby encouraging discussion. Through an expert-informed but peer-led model, the Functional Evidence Discussion Forum supports discussion-based, collaborative assessment of functional data for application in clinical curation. Importantly, our results indicate an increase in comfort in functional evidence use by participants through participation (Figure 5A and 5B). While we saw an initial drop in confidence of participants within the program, we attributed this to the previously reported effect of unconscious incompetence ^22^, and importantly note an increase in confidence across the span of the program (Figure 5C). Acknowledging that many factors impact day-to-day practice, as a future exercise it would be extremely informative to determine the moderate or even long-term impact of this program on variant classification outcomes and patient management.

These results build on previous results from the AusMAVE Education Working group and collaborators, calling internationally for expert recommendations to improve functional data evaluation ^8; 10; 23^. Previous work indicated that Australia followed a process generally aligned to the ClinGen variant classification recommendations ^8–10^.The workshops delivered content which responded to lower stakeholder comfort with high-throughput assays. The consultation results generated within the training program identified why and what aspects of expert advice may provide most benefit. In our training program we presented the current recommendations for functional evidence evaluation^3^ and saw that: initially there was poor knowledge, understanding and application of these recommendations; during a workshop exercise the recommendations were not applied consistently, despite prior detailed description of calibration processes in the same workshop (Figure 2). Variation in functional data evaluation was exacerbated by specific areas of difficulty including uncertainty in assessing model system appropriateness, conflicting assay results, and controls selection. The participants identified barriers aligned with their preferences on topic discussion and for continued learning, with appropriate disease models and conflicting data being among the topics of highest participant interest, see Figure 4C. identified previously through a needs survey ^8^. As participants became more comfortable with current recommended processes for calibration of higher throughput data, in Workshop 2, we noted high interest from the participants in tools and expert recommendations relevant to application of these types of assays (Figure 3). While some of the lack of comfort in functional evidence evaluation stemmed from lack of knowledge, the results indicate that another component was lack of confidence in the assay applicability and appropriateness, implying that assays are not being conducted or reported to a standard required for use as clinical evidence. We suggest this may underpin the identified preference for expert advice, and that practice support, along with knowledge development, may better resolve the lack of comfort in functional evidence evaluation and application. Additionally, the results clarified potential future approaches to improving practice comfort, as we identified difficulty in evaluating appropriateness of model and approach to resolving conflict as priority areas for practice development and support. Focusing on these priority areas are likely to result in the highest return for improved practice development and more confident use of functional evidence in clinical diagnostic practice.

An evolving and practical training program is required to support the rapidly developing field of functional evidence evaluation. We present our model and the associated resources for use by others in the development of training in functional evidence use. We recommend stakeholder and end-user co-design to maximise engagement and inform an approach with best results. We recommend using a well-established framework for training program development, in this case we used the Program Logic Model ^14^. We also recommend integrating evaluation planning from the initial outset of the project ^13^. Our program surveyed comfort with functional evidence evaluation at multiple time points, the results of which suggest success in improving comfort. Lastly, we recommend that implementing community-based discussion, involving members across field and expertise levels, will ultimately enable capability building in functional evidence evaluation practice that reflects stakeholder’s needs and preferences, and is responsive to the evolving practice.

## Data Availability

All data produced in the present work are contained in the manuscript or are available online at https://zenodo.org/communities/ausmave

## Author Contributions (CRediT statement)

Conceptualization: RMV, BT, AFR, ABS

Methodology: RMV, BT, TM, AFR, ABS

Formal analysis: RMV, BT, ABS

Investigation: RMV, BAS, ABS

Resources: RMV, ABS

Data Curation: RMV

Writing - Original Draft: RMV, BT, ABS

Writing - Review & Editing: RMV, BT, ET, CC, BAT, LS, CNH, BL, EPM, JC, AFR, ABS

Visualization: RMV

Supervision: RMV, ABS

Project administration: RMV, AFR, ABS

Funding acquisition: AFR, ABS

## Acknowledgements

A great many thanks to the Australian, New Zealand and International Functional Evidence community members that have contributed to the larger consultation and participated in the described workshops. Thanks also to the Atlas of Variant Effect community that have been instrumental in the development of this program and supporting the continuing use of the program outcomes. While it would be difficult to identify all individually, the entire international community has been overwhelmingly generous in contributing their opinion and expertise to this work.

## Ethics Declaration

All research undertaken in this study was approved by the Human Research Ethics Committee of QIMR Berghofer, Project ID P3920.

