## Supplemental Information for "Community co-designed workshops build confidence in use of functional evidence for variant classification"

#### Index

AusMAVE Education Project - Program Summary .... 3
Background .... 3
Workshop 1 .... 3
Objectives .... 3
Participants .... 4
Content .... 4
Consultation .... 5
Outcomes .... 5
Recommendations .... 9
Summary .... 10
Workshop 2 .... 10
Objectives .... 10
Participants .... 10
Content .... 13
Consultation .... 14
Outcomes .... 14
Recommendations .... 15
Summary .... 16
Functional Evidence Activities .... 16
Objectives .... 16
Participants .... 16
Content .... 17
Consultation .... 17
Outcomes .... 17
Recommendations .... 17
Summary .... 17
Workshop 3 – Discussion Forum Launch Oct 2024 .... 18
Objectives .... 18
Participants .... 18

Content ........ 19
Consultation ........ 19
Outcomes ........ 19
Recommendations ........ 21
Summary ........ 21
References ........ 21

### AusMAVE Education Project - Program Summary

#### Background

The AusMAVE Education program was originally informed by a survey of Australian diagnostic genomics professionals [1]. The survey assessed participants' current practice, barriers and facilitators for the purpose of improving the application of functional evidence in variant classification.  The program was then developed with support of an advisory committee, the AusMAVE Education Working Group, and in consultation with the functional evidence community. Associated program documents can be accessed through Zenodo via the AusMAVE project webpage, www.qimrb.edu.au/ausmave.

#### Workshop 1

In-person 17th November 2023

This in-person workshop was delivered at the Human Genetics Society of Australasia (HGSA) https://hgsa.org.au/, Australian Society of Diagnostic Genomics (ASDG) Special Interest Group (SIG) day meeting in Melbourne Australia, 17 November 2023. The expert advisory group identified that applied scientists performing variant classification were the program's primary target participant group, so Workshop 1 participants were recruited by directly targeting the already established society meeting,

**Objectives**

- To engage potentially relevant audience with the value and benefit of functional evidence

- To identify sources of functional evidence for use in variant curation
- To provide introductory approaches on assessing functional evidence quality
- To elicit feedback about current approaches to functional evidence in variant curation
- To identify opportunities to support users and potential users of functional evidence

**Participants**

Intended: HGSA members, diagnostic laboratory scientists and researchers, delivered at the November 2023 ASDG SIG meeting.

38 workshop attendees participated in the consultation, all members of the Australian Society of Diagnostic Genomics (ASDG).

**Content**

Slides: https://doi.org/10.5281/zenodo.13129941

Structure:

| Time frame | Topic | Details | Poll |
|---|---|---|---|
| NA | Enable completion of the Needs survey prior to workshop | | |
| 5 min | Problem and opportunity | Terminology used<br>Inconsistent functional evidence use (Shariant)<br>MAVE data increasing (MAVE DB) | |
| 5 min | Introduction to functional evidence and its use | 1)What is functional evidence<br>2)Low vs High throughput | |

| | | | |
|---|---|---|---|
| 5 min | Recommendations for functional evidence use | Present the current best recommendations and (MAVE) | Comments on Brnich, what questions do you have? |
| 5 min | Case study - introduce | Recommendations for how to find functional data. (ie MAVE DB) - case study as example | What functional data can you find? How did you find it? |
| 10 min | Case study - Where to find functional evidence | Low throughput assay - how to assess if class is appropriate?<br>How to calculate the strength - case study as example | How would you assess this assay, can the class be used? What evidence strength would you apply? |
| 5 min | Case study | High throughput - how to assess if appropriate (cheat with VCEP)<br>How to read the scores - case study as example | How would you assess this assay?<br>pos/neg controls, validation data |
| 5 min | Where to find out more | Present a list of resources for learning and/or using | Does anyone have learning resources/use resources to share? Ask if the audience knows/uses the resources presented |
| 5 min | Questions/ discussion | Ask participants their preferences for future education / resources / other activities. | |
| NA | Leave participants with a resource pack<br>Finish with an exit survey. | | |

**Consultation**

Consultation survey: DOI: 10.5281/zenodo.17532728

**Outcomes**

Workshop report: https://doi.org/10.5281/zenodo.17538260

Workshop 1 – supplementary results

Supplemental results indicating Workshop 1 participant opinion on evidence assessment when considering Assay 1, the gamma irradiation sensitivity assay in HCC1973 from Scully et al. 1999 PMID:10635334 [2].

Is the assay class an appropriate model of disease?

| Appropriate | # participants | % participants |
|---|---|---|
| Yes | 26 | 78.8 |
| Unsure | 6 | 18.2 |
| No | 1 | 3.0 |

Is this specific assay valid for use?

| Valid | # participants | % participants |
|---|---|---|
| Yes | 26 | 78.8 |
| Unsure | 5 | 15.2 |
| No | 2 | 6.1 |

What functional evidence (direction) could you apply?

| Strength | # participants | % participants |
|---|---|---|
| Pathogenic | 23 | 69.7 |
| Unsure | 10 | 30.3 |

What functional evidence (strength) could you apply?

| Direction | # participants | % participants |
|---|---|---|
| Moderate | 4 | 12.1 |
| Supporting | 18 | 54.5 |
| Unsure | 11 | 33.3 |

Participant opinion on evidence evaluation considering Assay 2, the High-throughput (SGE) SNV competition assay in HAP1 cells in Findlay et al. 2018 PMID:30209399 [3].

Can you calculate Odds Path?

| Odds Path | # participants | % participants |
|---|---|---|

| | | |
|---|---|---|
| Yes | 27 | 81.8 |
| Unsure | 5 | 15.2 |
| No | 1 | 3.0 |

What functional evidence (strength/direction) could you apply?

| Strength | # participants | % participants |
|---|---|---|
| Strong | 14 | 42.4 |
| Moderate | 5 | 15.2 |
| Supporting | 3 | 9.1 |
| Unsure | 11 | 33.3 |

The activity performed in the Functional Evidence Workshop shows variation in how expert diagnostic scientists find, consider and apply functional evidence in variant curation.

Curators identified that sourcing functional evidence for a specific variant was an area of difficulty. They generally sourced data for functional evidence through the medical literature, mastermind, ClinVar and google (in that order).

The workshop suggested that curators require improved methods for identifying functional evidence for a specific variant.

<u>Post-workshop assessment</u>

Participants were requested to fill and options post-workshop assessment, intended to survey the workshop content. The following results present the post-workshop assessment results for n = 6 workshop participants.

The learning outcomes (objectives) were relevant to the goal of improving the use of functio nal evidence

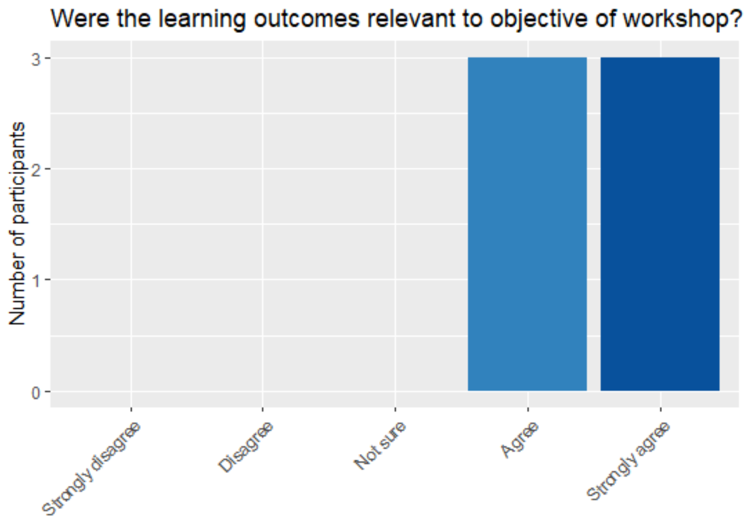
Were the learning outcomes relevant to objective of workshop?
Number of participants
0
1
2
3
Strongly disagree
Disagree
Not sure
Agree
Strongly agree

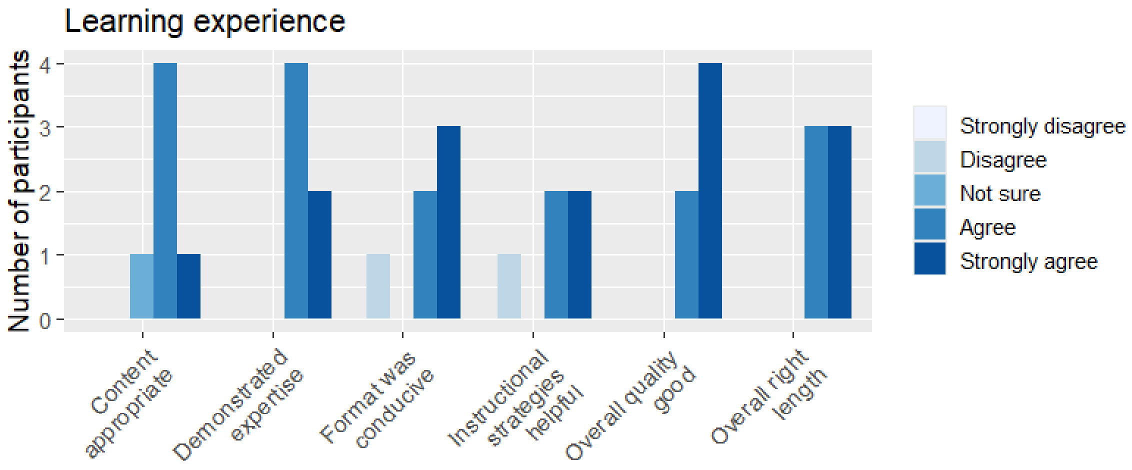
Learning experience
Number of participants
0
1
2
3
4
Content appropriate
Demonstrated expertise
Format was conducive
Instructional strategies helpful
Overall quality good
Overall right length
Strongly disagree
Disagree
Not sure
Agree
Strongly agree

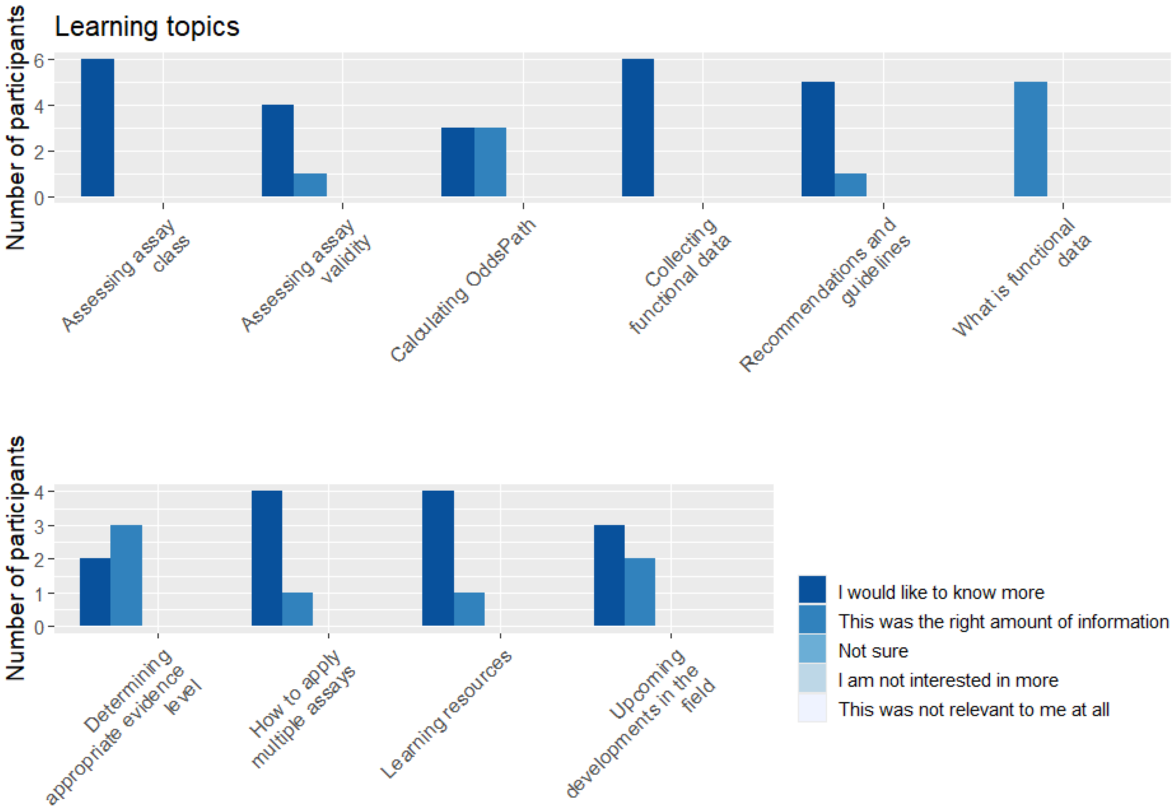

**Recommendations**

Based on the results of the workshop 1 activity, the MAVE Education project drafted the following aims to:

Collate and share information resources on use of functional evidence in variant classification, including expert recommendations.

Investigate the best methods for identifying functional data relevant to a specific variant.

Investigate the concept of assay class, with an aim to build recommendations for assay evaluation.

Align these activities with ACMG/AMP recommendations as per (Brnich et al., 2019; Richards et al., 2015)[4,5].

Legacy resources would support sustained use of the content.

Case-based and peer-supported training is preferred.

Workshop attendees identified that the application of functional evidence in their practice was impacted by practical reasons including time limitations for functional evidence assessment, time limitations for training in functional evidence assessment and detailed searching and assessment being outside of the scope of curator’s roles.

**Summary**

Workshop 1 indicated high interest in the topic from the diagnostic genomics community. It supported focussing on the diagnostic genomics and variant classification community as a primary stakeholder.

#### Workshop 2

Online workshop, 19 June 2024.

**Objectives**

- To provide hands-on workshop on using experimental data in variant classification.
- Support evaluating functional evidence according to Brnich et al.[4] and Gelman et al.[5].
- To capture the thought logic of curation scientists evaluating functional evidence in ‘real case’ scenarios
- To survey knowledge level of participants

**Participants**

Intended: Diagnostic Scientists, or any applied genetics specialist that is involved in curation or engaging with functional evidence.

Registration: 104 registrations total

Attendance: 80 logged in on the day (some laboratories attended as a group)

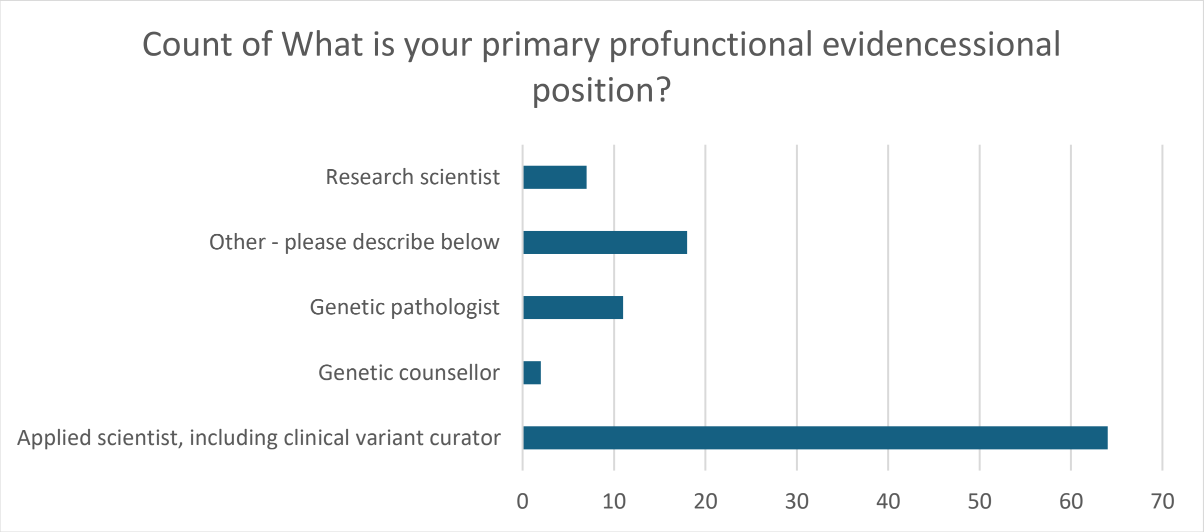

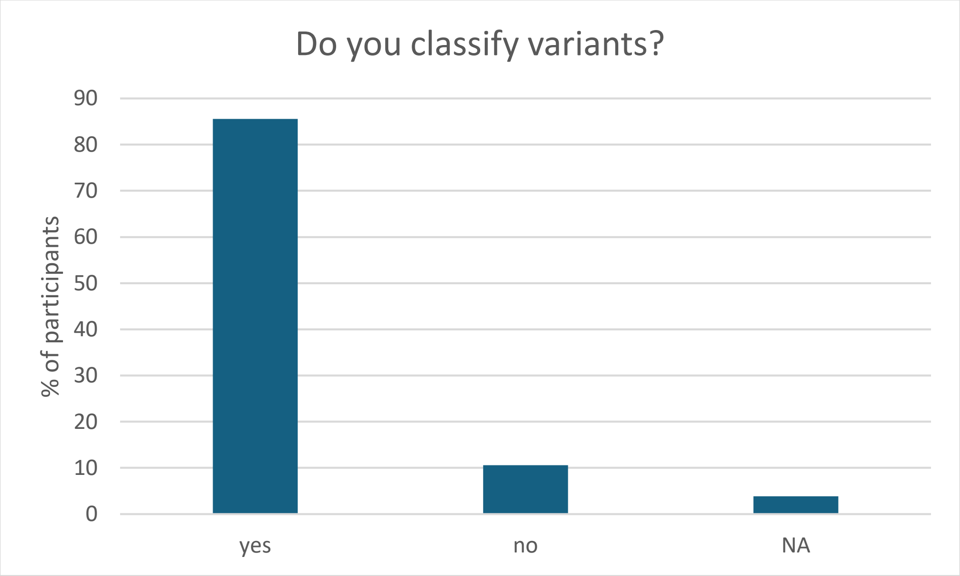

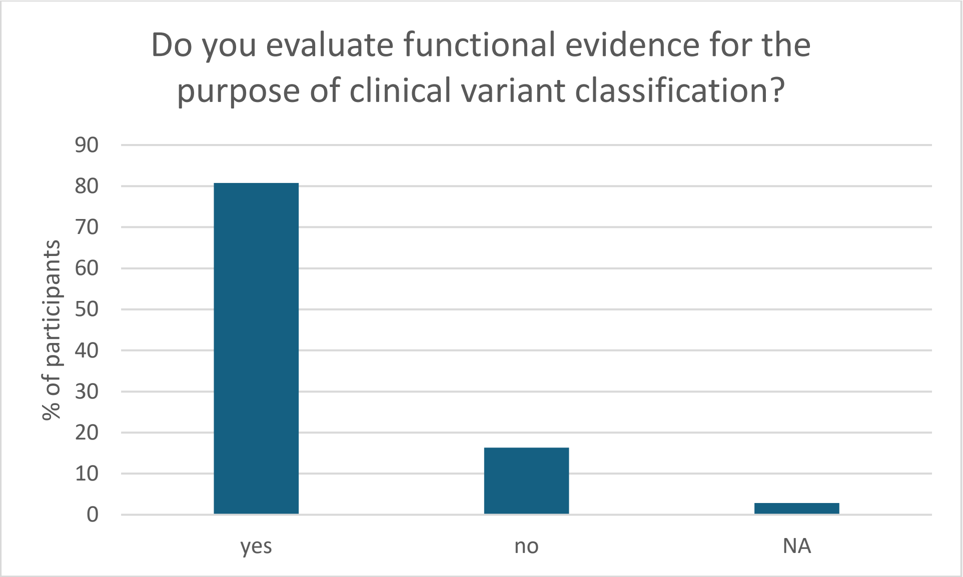
Do you evaluate functional evidence for the purpose of clinical variant classification?
% of participants
90
80
70
60
50
40
30
20
10
0
yes
no
NA

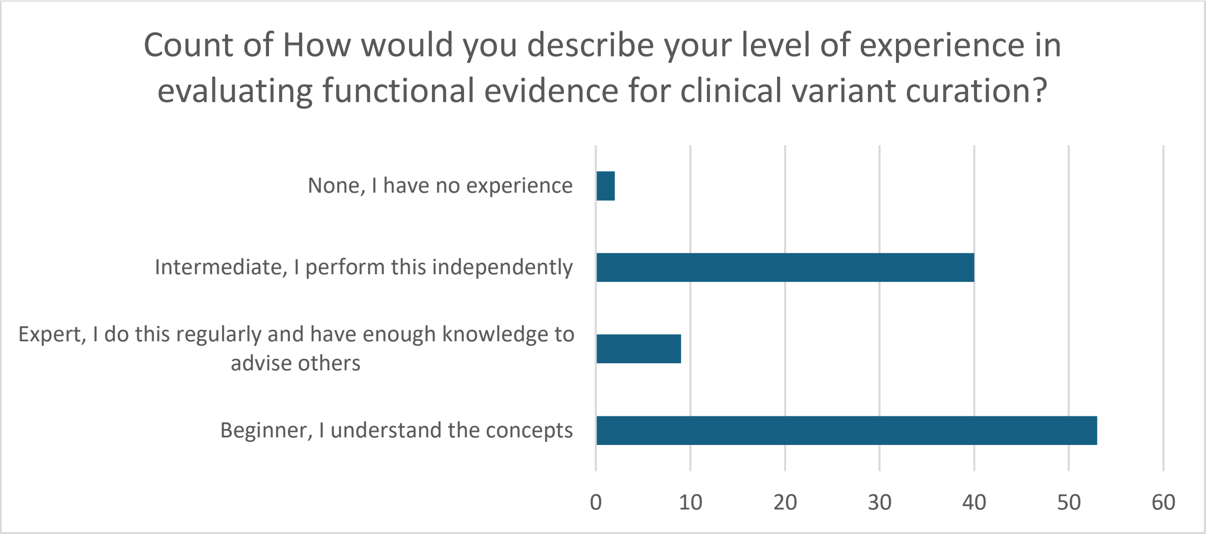
Count of How would you describe your level of experience in evaluating functional evidence for clinical variant curation?
None, I have no experience
Intermediate, I perform this independently
Expert, I do this regularly and have enough knowledge to advise others
Beginner, I understand the concepts
0
10
20
30
40
50
60

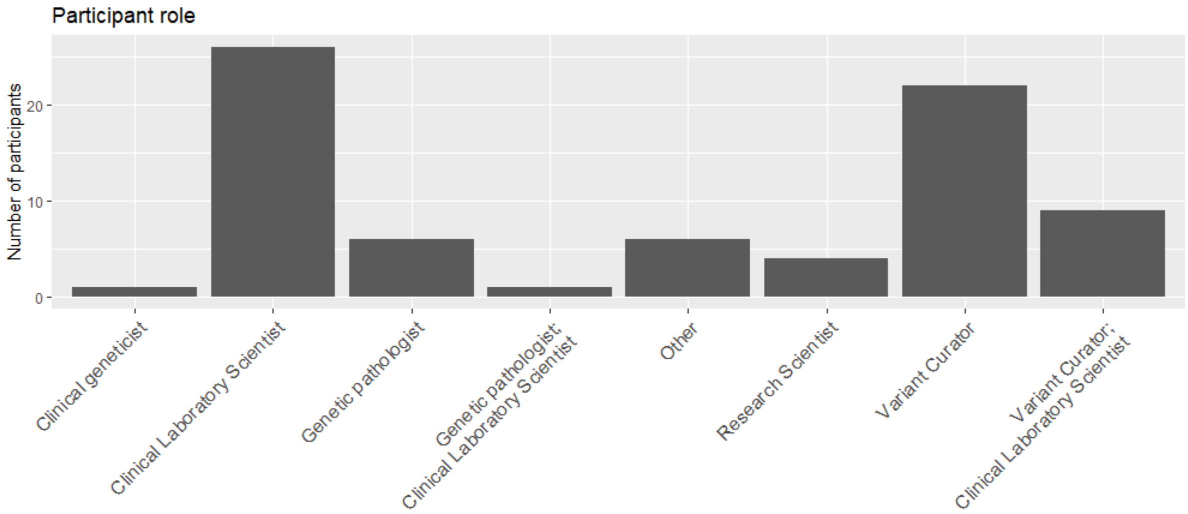

**Content**

Slides: Slides: https://doi.org/10.5281/zenodo.13131424

Structure:

| | | |
|---|---|---|
| 5 min | Project introduction | Explanation on the research activity |
| 30 min | Introduction on functional evidence. | Information session aimed at going over Brnich and Gelman<br>Anonymous poll of questions from the audience (questions about functional evidence evaluation) |
| 10 min | Case example | Led by demonstrator, using a single case example (similar to workshop 1) |
| 20 min | Case studies (breakout groups) | From an established series of case studies, allocate one per break out group<br>Break-out groups independently discuss and evaluate the case given<br>Case study evaluation outcomes captured by form (analyse results and variation) |
| 30min | Case Presentations | (6 x 5min) Presentation of break out group cases (5min each assuming 6 break out groups)<br>One of the group members presents a summary of their evaluation |
| 10 min | Open Discussion and Q&A | Discussion on Importance/significance Challenges<br>Discussion on Best Practices |
| 10 min | Conclusion and Wrap-Up | Summary of key concepts and techniques covered during the workshop |

**Consultation**

Workshop 2 presented demonstrator case studies based on an assay evaluation scenario. Discussion was invited on the scenarios and opinion on mechanisms to support practice were asked that aligned with the case studies. Opinion on OddsPath/LR calculation was specifically asked.

Consultation questions: https://doi.org/10.5281/zenodo.17461665

**Outcomes**

Workshop report: https://doi.org/10.5281/zenodo.17538303

Polls established the demographics of participants, asked regarding comfort in calibration and evidence strength calculation and polled opinion on clinical controls and logic on evidence evaluation.

Polls available on Zenodo: Villani, R., Spurdle, A., & Rubin, A. (2024). Workshop Polls: Functional evidence evaluation workshop, June 2024 (2024.06.19.1). Zenodo. https://doi.org/10.5281/zenodo.17461665

Workshop 2 identified a building knowledge of OddsPath/LR, but also participants encouraged support in practice building in the area. 98% of the participants indicated they would like to access the activities afterward.

In the workshop we also initiated consultation around continuing practice and learning support with the participants, and identified a strong support for a continuing discussion forum.

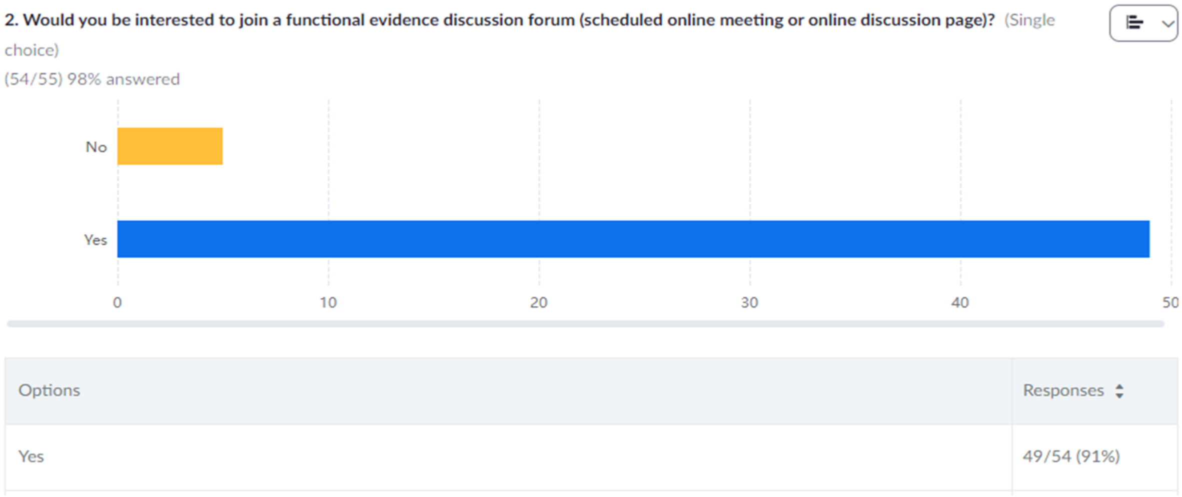

| Options | Responses |
|---|---|
| Yes | 49/54 (91%) |

5 online activities were shared at the workshop to provoke discussion. The activities, based on participant opinion, were converted to legacy activities, see below.

Clinical consultants (specialist medical doctors) were engaging with functional data/assays outside of the formal classification/test report. Their assessment of functional data was often a catalyst for reanalysis or continuing diagnostic testing/analysis.

A likelihood calculator spreadsheet was shared in the meeting that was well received, and highlighted the need for publicly available resources and tools to support evidence strength calculation.

**Recommendations**

To provide tools that facilitate functional evidence evaluation practice, while concurrently supporting learning and minimising other identified barriers, such as lack of time.

Enable variant classification scientists to collate, share and use the currently available recommendations and tools.

To build a community of practice through an ongoing community based forum for discussion, supporting discussion with functional evidence community members around specific assays, assessment process, and application.

**Summary**

Based on the outcomes of Workshop 2, the workshop activities were converted to online, self-directed activities with answers supplied, see next section. In addition, this activity identified an interest in a continuing, peer-based approach to learning. Areas of particular interest for support were identified, namely conflicting evidence, appropriateness of assay systems, and how to select clinical controls to be used in clinical calibration.

#### Functional Evidence Activities

**Objectives**

- To provide legacy resources for case-based practice, as requested in Workshop 2.
- To gauge variation in the assessment of functional assays for use as functional evidence.

**Participants**

Intended: Distributed was originally limited to 104 Workshop 2 participants, all users of functional evidence, including diagnostic variant classification scientists, genetic pathologists and researchers.

Actual: Functional Evidence Evaluation Activity Instructional Slides are now available via Zenodo, link below, 77 views at 29.10.2025

**Content**

Instructional Slides: https://doi.org/10.5281/zenodo.13888494

4 independent online activities. Each presents a scenario for evaluating a functional assay for use in variant classification.

**Consultation**

Activity participants submit their opinion on the activity results, the integrated consultation can be found within the activity outcomes.

**Outcomes**

The Workshop 2 activities were converted to the described online activities, the results of the activities generated in the workshop were collected from groups, and were used only to develop the activities for public release.

**Recommendations**

Variant classification scientists want practice activities to support learning. These should be case and practice based.

**Summary**

The activities were used as a demonstrator for practice, however could be used as self-directed learning resources by the discussion forum members.

#### Workshop 3 – Discussion Forum Launch Oct 2024

Discussion Forum Launch, October 2024.

**Objectives**

To consult on the delivery of a Functional Evidence Discussion Forum, based on recommendations from Workshop 2. Aim to articulate specific approach to ensure value to the stakeholders.

**Participants**

Intended: Aimed at users of functional evidence, applied scientists, genetic pathologists and functional genomics researchers.

Actual: 76 people registered for the functional evidence forum, with 78 participants signing in on the day.

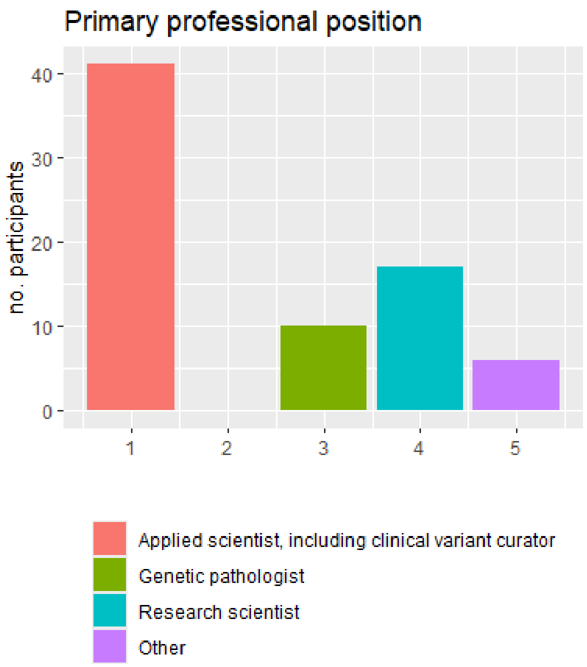

**Content**

Presentation 1: Dr Alan Rubin presented "*MaveDB: An international community database for functional assay data*". Slides were shared with the participants post the session.

Presentation 2: The Functional Evidence Discussion Forum Consultation

Structure:

| | | |
|---|---|---|
| 5 min | Project introduction | Explanation on the research activity and proposal to launch the forum |
| 20 min | Dr Alan Rubin presented "*MaveDB: An international community database for functional assay data*". | A session presenting the application and underlying concepts behind MaveDB. |
| 10 min | Open Discussion and Q&A | Led by facilitator |
| 25 min | Consultation survey | Identifying what the participants want and need from a forum to inform the continuing approach. |
| 5 min | Conclusion | |

**Consultation**

Workshop 3 was designed to determine if there was interest in the community for such an activity. Consultation focussed on determining the approach for the proposed continuing Functional Evidence Discussion Forum.

Consultation questions: https://doi.org/10.5281/zenodo.17538156

**Outcomes**

Workshop report: https://doi.org/10.5281/zenodo.17538345

To host 4 online discussion forum sessions and deliver a pilot online web-forum for 'out-of-session' discussion. We will aim to invite one researcher to present their assay and one curator to review a published assay at each session. The Discussion Forum will be reviewed at the end of 2025 to determine the plan for/if continuing.

1. Excellent interest with high participation rate, >70 participants online.
2. Curators are highly represented in the participant group (most interest/relevance for this group), however a range of other expertise was present/interested including pathology and research professionals. Should likely target to curators, but will give opportunity for researchers to collaborate.
3. Confidence for functional evidence evaluation was middle of the 1-5 range, most participants indicated 3 (5 being the highest comfort level).
4. Most participants supported a quarterly online seminar.
5. An online forum (potentially a discussion page) could be useful, and initiating a low impact option may be worth trialling. The continuation on this might be dependent on the engagement.
6. A wide range of topics and presentation types are useful.
7. Topics of interest to highest number of people were 'Evaluating assays as an appropriate model for disease' and 'Can I 'use' this assay? (Curator presenting).
8. Discussing public/published data would be no problem. These conditions are perfectly suitable for the Functional Evidence Discussion Forum.
9. We should recruit from the participants to nominate assays and to present them for discussion, this will identify most useful discussions but also support sharing knowledge.
10. General support was indicated for recording and sharing, suggest establishing consent for recording sharing from the presenter on a case-by-case basis is appropriate.

**Recommendations**

- We should draft 'rules' for the Functional Evidence Discussion Forum
- Generally speaking it was agreed that NO patient information could be shared, but variant and phenotype information sharing would be fine.
- Suggest building a template for presenting an assay for discussion.

**Summary**

Workshop 3 defined the approach for the continuing forum. There was excellent interest and attendance, and the AusMAVE Education project team aim to continue delivering project outcomes through the approach defined by the stakeholders.
